# Effect of the 2025 National Institutes of Health grants disruption on first-time and mechanism-first principal investigators: a cohort study of 80,976 active awards

**DOI:** 10.64898/2026.05.22.26353911

**Authors:** Fares Alahdab, Bettina Mittendorfer

**Author notes:** **Corresponding Author:** Fares Alahdab, MD, MS, MSc, FAHA, FACC, Associate Professor, Department of Biomedical Informatics, Biostatistics, and Medical Epidemiology, Department of Medicine, Division of Cardiovascular Medicine, Director of Academic Programs in Health Informatics, Core Faculty – Institute for Data Science and Informatics, University of Missouri School of Medicine, Columbia, MO, USA, E.

## Abstract

**Objective:** To estimate the adjusted relative risk (RR) of administrative grant disruption faced by first-time and mechanism-first principal investigators (PIs) during the 2025 National Institutes of Health (NIH) grant disruptions.

**Design:** Retrospective cohort study linking NIH RePORTER data to a publicly curated registry of grants disrupted in 2025.

**Setting:** All NIH active research grants in fiscal years 2024 to 2025.

**Participants:** 80,976 active projects: 4,961 disrupted during the wave that peaked in May 2025, 76,015 non-disrupted controls.

**Main outcome measures:** Adjusted RR of disruption by two pre-specified first-time PI constructs: absolute first-time PI (no prior NIH grant) and mechanism-first PI (no prior NIH grant with the same activity code). Modified Poisson regression with institution-clustered standard errors adjusted for project, institutional, and geographic covariates. A pre-specified fiscal year 2024 common-anchor analysis addressed year-of-disruption confounding.

**Results:** Of 4,961 disrupted grants, 237 (4.8%) had an absolute first-time PI and 396 (8.0%) had a mechanism-first PI. After adjustment, absolute first-time PIs faced 77% elevated risk of disruption (RR 1.77, 95% CI 1.34 to 2.32) and mechanism-first PIs faced 57% elevated risk (RR 1.57, 1.16 to 2.11). Under the common-anchor analysis, the absolute first-time effect attenuated to RR 1.22 (0.95 to 1.58); the mechanism-first effect persisted (RR 1.48, 1.07 to 2.06). The elevated risk was concentrated in research-mechanism grants (RR 1.78, 1.26 to 2.52) and was robust across 8 of 9 pre-specified sensitivity analyses. The Track A start-time construct, which asks whether the disrupted project was the PI’s debut grant, yielded null estimates (RR 0.98, 0.93 to 1.04), with any effect concentrated entirely in newly started projects.

**Conclusions:** First-time and mechanism-first PIs faced disproportionately elevated risk of disruption during the 2025 NIH wave, concentrated in research-mechanism grants and robust to year-confounding-free identification. The relevant exposure was being early-career at the moment of administrative action, not at project initiation. The findings have direct implications for workforce equity in US biomedical research.

**Key messages:** *What is already known on this topic:* The 2025 National Institutes of Health grant disruption wave affected approximately 5,500 active awards across termination, freezing, and reinstatement statuses, with documented disproportionate impact on certain research areas, organisational types, and geographic regions. Whether the wave disproportionately affected investigators at specific career stages, particularly first-time principal investigators (PI), has not been formally analyzed using adjusted estimates from the full NIH grant universe. Methodological choices in defining first-time PI status, including the distinction between start-time and index-time exposures and the confounding role of project lifecycle stage, have not been systematically addressed in workforce analyses of the 2025 disruption.

*What this study adds:* After adjustment for mechanism, NIH institute, log cost, project age, support year, application type, institution type, and US region, absolute first-time PIs faced a 77% elevated relative risk of disruption (RR 1.77, 95% CI 1.34 to 2.32), and mechanism-first PIs faced a 57% elevated risk (RR 1.57, 1.16 to 2.11). The signal persists in a pre-specified fiscal year 2024 common-anchor analysis designed to eliminate year-of-disruption confounding, with a mechanism-first relative risk of 1.48 (1.07 to 2.06). The elevated risk is concentrated in research-mechanism grants and is robust across eight of nine pre-specified sensitivity analyses, indicating that administrative disruption fell disproportionately on the most vulnerable career stage in the biomedical research workforce. The risk reflects PI status at the calendar moment of disruption (index-time), not at project initiation (start-time), a distinction with direct implications for how workforce vulnerability is operationalised in future analyses of funding shocks.

## Introduction

In early 2025, the National Institutes of Health (NIH), the largest funder of biomedical research in the world, experienced a wave of administrative grant disruptions of unprecedented scale and pace. Between February 2025 and February 2026, peaking in May 2025, approximately 5,500 active research awards were terminated, frozen, or otherwise disrupted, representing committed federal research funding totaling several billion dollars [1]. Initial descriptive characterisations of the wave, drawing on public databases and academic monitoring efforts, documented its concentration in Northeastern research universities, its over-representation of grants addressing health equity and disparities, and its uneven impact across specific NIH institutes and centers [2–5].

Less well understood is whether the wave disproportionately affected investigators at specific career stages. Early-career investigators, particularly those holding their first NIH research award, occupy a structurally vulnerable position in the biomedical workforce. Many depend on a single grant to sustain a nascent research program, lack the resource buffers of established laboratories, and have not yet accumulated the institutional and reputational capital that protects more experienced PIs from administrative shocks [6–9]. More broadly, scientific funding exhibits a Matthew effect, whereby modest early advantages compound over a career into substantial long-term differences in publications, recognition, and renewal probability [10,11]. An acute administrative shock that falls disproportionately on first-time and mechanism-first PIs may therefore have effects that extend well beyond the immediate dollar value of the disrupted awards, by foreclosing the early career milestones on which the long-arc Matthew effect depends [7,8,12]. The NIH has long recognised this vulnerability through its New Investigator and Early Stage Investigator policies, which provide modest protective adjustments in peer review for investigators who have not previously held a substantial independent award [13]. Whether the 2025 disruption wave compounded this structural vulnerability is an open empirical question with direct implications for the long-term composition of the biomedical research workforce.

Two methodological challenges complicate this question. First, first-time PI status admits at least two operationally distinct definitions. A PI may be classified as first-time on the basis of whether they held any prior NIH grant at the time of the disruption (an index-time construct), or on the basis of whether they held any prior NIH grant when the disrupted project was initiated (a start-time construct). These constructs measure different things and can diverge substantially in populations with heterogeneous project ages. Second, the disruption wave occurred within a specific calendar window with a limited temporal range, and the relationship between PI experience and project lifecycle is non-random. First-time PIs naturally cluster in newly started projects, and newly started projects naturally have less calendar-time exposure to the disruption wave. Failure to adjust for this lifecycle structure can produce misleading conclusions in either direction.

Using NIH RePORTER data covering all active NIH research grants in fiscal years 2024 and 2025, linked to a publicly curated registry of disrupted grants [1], we conducted a retrospective cohort study with two pre-specified exposure constructs and a primary analytic strategy designed to address lifecycle confounding. The primary outcome was administrative disruption during the wave, defined inclusively to capture all five disruption status categories present in the source registry.

We hypothesized a priori that first-time PIs would face elevated risk of disruption after adjustment for project lifecycle and other administrative covariates, and that the effect would be most pronounced in traditional research mechanisms where first-time PI status carries the greatest precarity.

## Methods

### Data sources

We used two data sources in linkage. The first was the NIH RePORTER data, which contains administrative metadata for all NIH-funded research projects, including project identifiers, principal investigator names, awarding institute or center, activity code, application type, support year, funding amount, organisational metadata (institution name, state, congressional district, institution type), and project start and end fiscal years. We used data covering the analytic window of fiscal years 2024 to 2025. Following standard procedures for the integration of NIH grant data, we used the core project number (a concatenation of the activity code, administering institute, and serial number) as the canonical project identifier, and the project’s first fiscal year of support as its start fiscal year.

The second source was a publicly curated registry of grants disrupted during the 2025 wave [1]. For each grant, the registry includes the full award number, status, and the relevant administrative event dates (termination date, freeze date, unfreeze date, and estimated reinstatement date, as applicable). We treated all five status categories as constituting the disrupted cohort, consistent with the source registry construction, and use the term “disrupted” throughout to refer to this composite outcome.

### Disruption date and temporal filter

For each disrupted project, we constructed a coalesced disruption date as the earliest event date available, in priority order: termination date, freeze date, then unfreeze date. The coalesced disruption date was populated for 99.9% of disrupted fiscal year rows in the analytic cohort.

### Cohort construction and active window

The active window was defined as fiscal year 2024 to fiscal year 2025. For each project, we constructed a single index fiscal year. For disrupted projects, the index fiscal year was the latest fiscal year of support within the active window, representing the calendar context of the disruption. For non-disrupted projects, the index fiscal year was the earliest fiscal year of support within the active window, representing the analogous active period. We excluded projects with missing principal investigator data after PI identifier normalization.

### Exposure definitions

We defined two pre-specified exposure constructs.

The primary (index-time, Track B) exposure was constructed using each project’s PI history up to and including the project’s index fiscal year. A PI was classified as absolute first-time at index if they had zero prior NIH grants of any kind starting in any fiscal year before the index fiscal year, and as mechanism-first at index if they had zero prior NIH grants in the same activity code (e.g., R01) starting before the index fiscal year. These constructs capture whether the PI was new to NIH or new to the activity-specific mechanism at the calendar moment of the disruption.

The secondary (start-time, Track A) exposure was constructed using each PI’s history up to and including the project’s start fiscal year. A PI was classified as absolute first-time at start if their earliest NIH grant began in the project’s start fiscal year, and as mechanism-first at start if their earliest NIH grant in the same activity code began in that fiscal year. These constructs capture whether the disrupted project was the PI’s debut grant in absolute or mechanism-specific terms.

The two tracks measure structurally different quantities. The same PI can be Track A first-time (no prior grant when the project started), but not Track B first-time (the PI accumulated additional grants by the time of disruption). In a stable cohort with no calendar-time exposure heterogeneity, the two tracks would converge after adjustment. When they diverge, the divergence is informative about the role of project lifecycle.

For sensitivity analyses, we additionally constructed a four-level mutually exclusive PI status variable (absolute first-time, mechanism-first only, experienced in mechanism, experienced across mechanisms) using the Track B index-time framing.

### Principal investigator identifier

The NIH RePORTER data includes the contact principal investigator’s name in semi-structured form, with role tags and multi-PI separators present in some records. We applied a strict normalization pipeline that stripped role tags and credentials, took the contact PI as the canonical PI for each grant, and consolidated formatting variants. Coverage of the resulting strict PI identifier in the full RePORTER universe was 99.7% of all fiscal year rows. As a sensitivity analysis, we additionally constructed a lenient PI table including all listed PIs from multi-PI grants and re-estimated the headline effect under this alternative linkage.

### Confounders

In the primary adjusted models we included the following pre-specified covariates: activity code (mechanism), administering institute or center, log of total cost (with a continuous specification in the primary model and a categorical specification as sensitivity), project age band at index (0 [new], 1, 2 to 3, 4 to 5, or greater than 5 years since project start), support year, application type, institution type with an explicit missing indicator because approximately 28% of records have missing institution type, and US region (Northeast, Midwest, South, West, Territory).

### Statistical analysis

The primary estimator was a modified Poisson regression with a log link, estimated by generalized linear model with a Poisson family and robust (“HC1”) standard errors clustered at the institution (organization name) level [14,15]. This estimator yields valid relative risk estimates and confidence intervals for binary outcomes with substantial adjustment, and the cluster-robust standard errors account for the non-independence of grants within the same institution. We also estimated logistic regression as a complementary model.

For descriptive summarization, we used direct standardization to compute the observed-to-expected (O/E) ratio of first-time PI representation in the disrupted cohort relative to the non-disrupted cohort, within strata defined jointly by activity code, NIH institute, index fiscal year, cost band, and project age band [16]. Confidence intervals for the O/E ratio were estimated by a 1,000-iteration nonparametric bootstrap [17].

To address the structural asymmetry between the index fiscal years of disrupted projects (clustered in 2025) and non-disrupted projects (clustered in 2024), we conducted a pre-specified common-anchor analysis restricting both groups to fiscal year 2024 index records and re-estimating the primary Poisson model. This analysis is robust to confounding by year of disruption [18].

### Sensitivity analyses

We conducted nine pre-specified sensitivity analyses: S1 restricted to research mechanism codes (R, U, P, D, S); S2 restricted to R01 awards only; S3 used the four-level mutually exclusive PI status variable; S4a excluded grants flagged as reinstated; S4b descriptively compared first-time PI prevalence between reinstated and not-reinstated disrupted grants; S5 substituted the lenient PI linkage; S6a and S6b varied the cost adjustment specification; and S7 was a complete-case analysis dropping observations with missing institution type.

### Subgroup interactions

We pre-specified three subgroup interaction analyses: by mechanism category (research, training/fellowship, career development, other), by project age band, and by cost band. For the mechanism category and cost band, we used the Track B index-time absolute first-time exposure. For the project age band, we used the Track A start-time exposure because Track B is structurally degenerate for projects with an age greater than zero (a PI cannot be index-time first-time after their first year of support).

### Burden estimation

We report two complementary burden estimates. The gross burden is the count and total committed award value of disrupted grants in each first-time PI category. The adjusted-attributable burden applies the attributable fraction formula AF = (RR − 1) / RR to the count of disrupted first-time PI grants, with the corresponding cost calculated at the median award amount within the affected subgroup.

### Software and reproducibility

All analyses were conducted in Python 3 using pandas, NumPy, statsmodels, and scikit-learn.

### Reporting

This study is reported in accordance with the Strengthening the Reporting of Observational Studies in Epidemiology (STROBE) statement [19].

## Results

### Cohort

Of 5,474 grants in the source registry of disrupted awards, 5,440 were successfully matched to the RePORTER universe. After restriction to fiscal years 2024 to 2025 and exclusion of 220 projects with missing principal investigator data, the analytic cohort comprised 80,976 unique projects, of which 4,961 were disrupted and 76,015 served as non-disrupted active controls. A coalesced disruption date was populated for 99.9% of disrupted fiscal year rows. Figure 1 displays the cohort flow.

**Figure 1.**
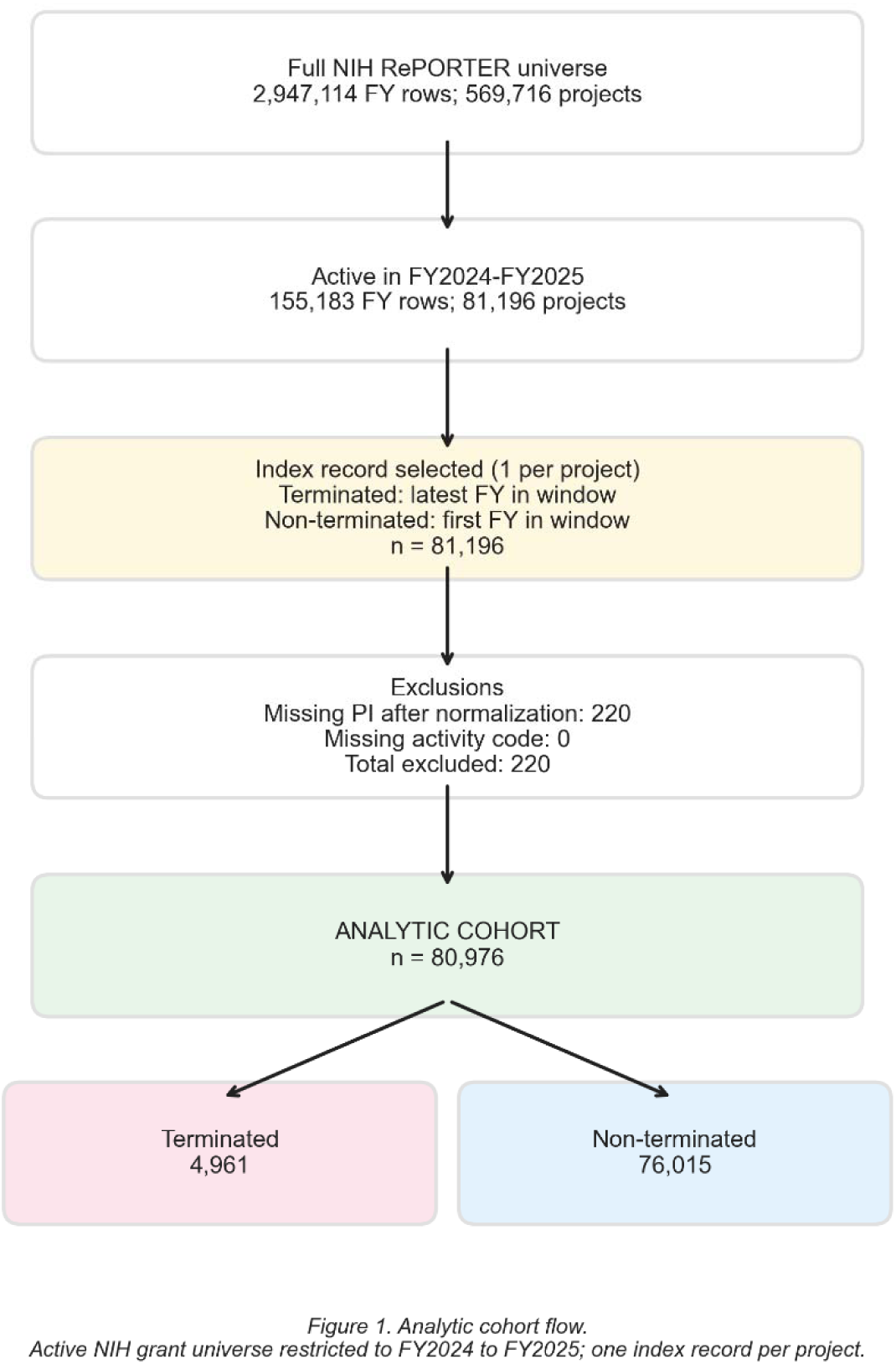
Cohort flow diagram. STROBE-style cohort flow showing the construction of the analytic cohort from the source NIH RePORTER universe (n=2,947,114 fiscal year rows on 569,716 projects) and the source registry of disrupted grants (n=5,474 grants). After restriction to the fiscal year 2024 to fiscal year 2025 active window, application of the coalesced disruption-date filter (1 February 2025 onward), and exclusion of records with missing principal investigator data, the final analytic cohort comprised 4,961 disrupted projects and 76,015 non-disrupted active controls.

Disrupted and non-disrupted grants differed substantively across most measured characteristics (Table 1). The disrupted cohort was geographically concentrated in the Northeast (48.2% of disrupted versus 27.7% of non-disrupted, p < 10^⁻^²□□), enriched for schools of medicine and schools of public health (57.1% and 9.3% versus 45.5% and 2.8%, respectively), and skewed toward more mature projects (median years since project start 2.0 versus 1.0, p < 10^⁻^¹□□). The four-level PI status distribution differed significantly between groups, with experienced PIs across multiple mechanisms over-represented among the disrupted (52.6% versus 33.7%) and mechanism-first-only PIs under-represented (3.2% versus 13.3%).

**Table 1.** Baseline characteristics of the analytic cohort. Selected baseline characteristics of the analytic cohort of 80,976 NIH active research grants in fiscal years 2024 to 2025, comparing the disrupted cohort (n=4,961) and the non-disrupted control cohort (n=76,015).

| Characteristic | Disrupted<br>(n=4,961) | Non-disrupted<br>(n=76,015) | p-value |
| --- | --- | --- | --- |
| PI status (Track B, index-time) |  |  |  |
| Absolute first-time | 237 (4.8%) | 8,256 (10.9%) | $1.1 \times 10^{-1}$ □<sup>1</sup> |
| Mechanism first-time | 396 (8.0%) | 18,336 (24.1%) | $3.5 \times 10^{-1}$ □ □ |
| PI status (Track A, start-time) |  |  |  |
| Absolute first-time at start | 1,495 (30.1%) | 21,239 (27.9%) | $9.1 \times 10^{-}$ □ |
| Mechanism first-time at start | 3,330 (67.1%) | 49,101 (64.6%) | $3.2 \times 10^{-}$ □ |
| PI status (4-level) | | | $5.9 \times 10^{-221}$ |
| Absolute first-time | 237 (4.8%) | 8,256 (10.9%) |  |
| Mechanism first-time only | 159 (3.2%) | 10,080 (13.3%) |  |
| Experienced in mechanism | 1,954 (39.4%) | 32,024 (42.1%) |  |
| Experienced across mechanisms | 2,611 (52.6%) | 25,655 (33.7%) |  |
| Project lifecycle |  |  |  |
| Median years since project start | 2.0 [1.0, 4.0] | 1.0 [0.0, 3.0] | $<10^{-1}$ □ □ |
| Age band 0 (new) | 503 (10.1%) | 27,562 (36.3%) |  |
| Age band 1 | 1,149 (23.2%) | 12,416 (16.3%) |  |
| Age band 2-3 | 1,646 (33.2%) | 17,561 (23.1%) |  |
| Age band 4-5 | 884 (17.8%) | 8,027 (10.6%) |  |
| Age band >5 | 779 (15.7%) | 10,449 (13.7%) |  |
| US region | | | $5.7 \times 10^{-2}$ □ □ |
| Northeast | 2,389 (48.2%) | 21,075 (27.7%) |  |
| Midwest | 937 (18.9%) | 13,903 (18.3%) |  |
| South | 714 (14.4%) | 20,750 (27.3%) |  |
| West | 899 (18.1%) | 16,573 (21.8%) |  |
| Institution type (top categories) |  |  |  |
| Schools of medicine | 2,833 (57.1%) | 34,617 (45.5%) |  |
| Schools of public health | 462 (9.3%) | 2,131 (2.8%) |  |
| Schools of arts and sciences | 570 (11.5%) | 6,477 (8.5%) |  |
| Missing institution type | 297 (6.0%) | 22,424 (29.5%) |  |

*Values are n (%) or median [IQR]. p-values from chi-square tests for categorical variables and Mann-Whitney tests for continuous variables.*

### Track A and Track B exposure prevalences

Of the 4,961 disrupted grants, 1,495 (30.1%) had a Track A absolute first-time PI at the start of the disrupted project, and 3,330 (67.1%) had a Track A mechanism-first PI. In the non-disrupted control group, the corresponding proportions were 27.9% and 64.6%. By the Track B index-time framing, 237 disrupted grants (4.8%) had an absolute first-time PI and 396 (8.0%) had a mechanism-first PI, compared with 10.9% and 24.1% in the control group.

The crude unadjusted directions disagreed between tracks: Track A showed modest over-representation of first-time PIs in the disrupted cohort, while Track B showed substantial under-representation. This divergence reflects the influence of project lifecycle, which we address explicitly in the subsequent analyses. Figure 2 displays the Track A versus Track B gap across activity codes.

**Figure 2.**
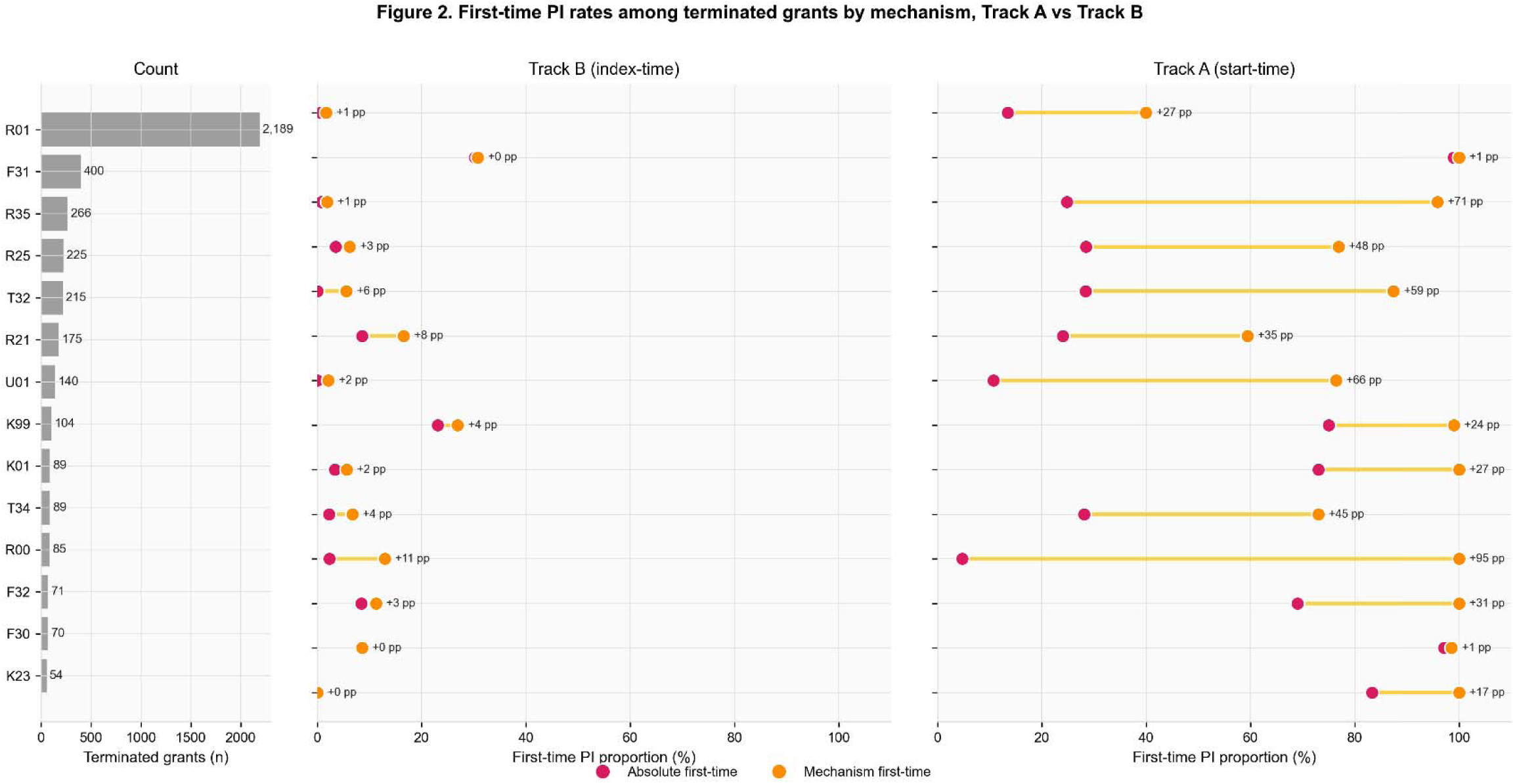
Mechanism gap between Track A start-time and Track B index-time first-time PI proportions. Dumbbell-style plot showing the gap between the Track A start-time proportion and the Track B index-time proportion of first-time PIs in the disrupted cohort, by activity code. The structural gap between the two reflects the influence of project lifecycle on the relationship between PI experience and exposure timing.

### Primary adjusted analysis

In the modified Poisson regression with institution-clustered standard errors and full covariate adjustment, Track B absolute first-time PIs faced a 77% elevated relative risk of disruption (adjusted RR 1.77, 95% CI 1.34 to 2.32, p=4.5x10⁻□). The effect for Track B mechanism-first PIs was a 57% elevated risk (adjusted RR 1.57, 1.16 to 2.11, p=0.003). The Track A start-time estimates were essentially null: absolute first-time at start RR 0.98 (0.93 to 1.04, p=0.52), and mechanism-first at start RR 1.05 (0.97 to 1.13, p=0.23). Figure 3 displays the O/E forest plot summarising the four primary estimates, and Table 2 provides the corresponding O/E summary.

**Figure 3.**
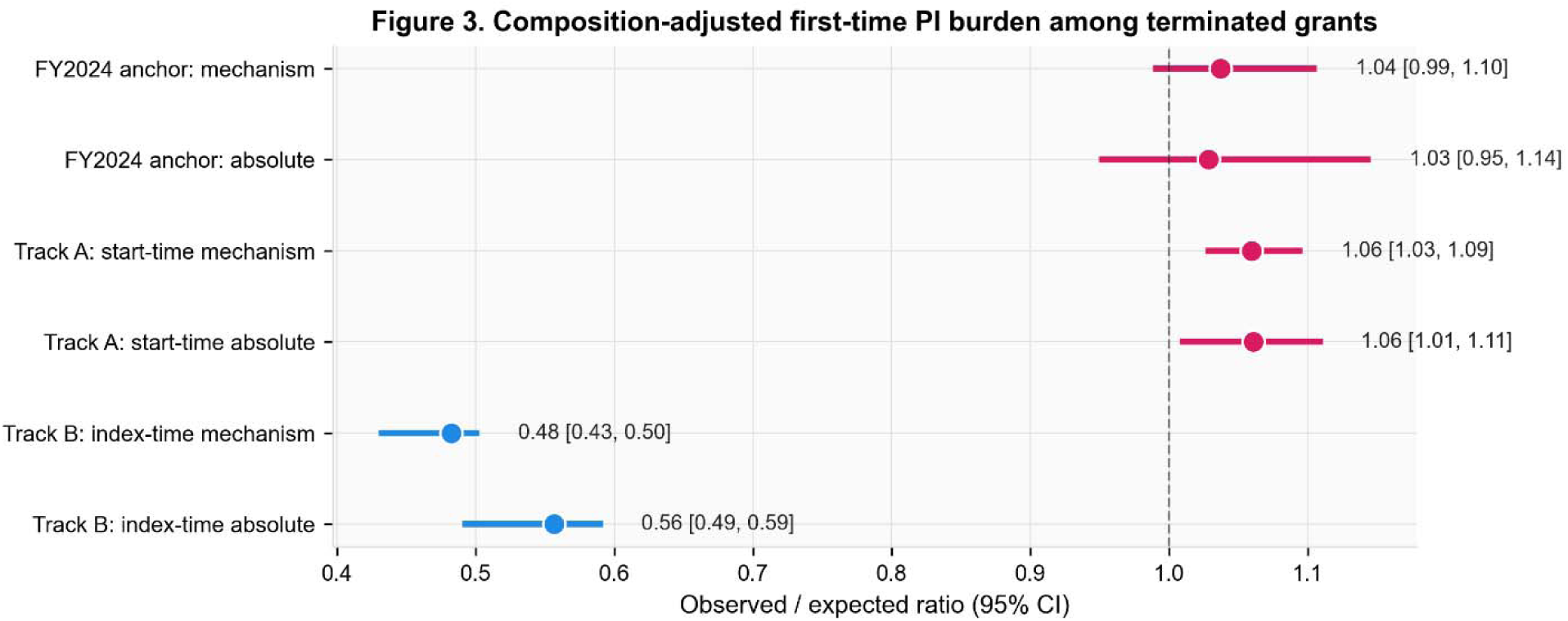
Observed-to-expected ratio forest plot for primary exposure constructs. Forest plot displaying the four primary O/E ratios with 95% bootstrap confidence intervals. Track B exposures (top two rows) yield O/E ratios below unity, indicating that within strata the disrupted cohort has fewer first-time PIs than expected from the non-disrupted distribution; this is the descriptive complement to the adjusted Poisson analysis (Results, Primary adjusted analysis). Track A exposures (bottom two rows) yield O/E ratios at or slightly above unity.

**Table 2.** Observed-to-expected (O/E) ratios for primary exposure constructs. Direct-standardized observed-to-expected ratios of first-time PI prevalence in the disrupted cohort relative to the non-disrupted cohort, within strata defined jointly by activity code, NIH institute, index fiscal year, cost band, and project age band. 95% confidence intervals from a 1,000-iteration nonparametric bootstrap.

| Exposure | Observed % | Expected % | O/E [95% CI] | Excess (pp) |
| --- | --- | --- | --- | --- |
| Track B: absolute first-time | 4.8% | 8.6% | 0.56 [0.49, 0.59] | −3.8 |
| Track B: mechanism first-time | 8.0% | 16.6% | 0.48 [0.43, 0.50] | −8.6 |
| Track A: absolute first-time at start | 30.1% | 28.4% | 1.06 [1.01, 1.11] | +1.7 |
| Track A: mechanism first-time at start | 67.1% | 63.3% | 1.06 [1.03, 1.09] | +3.8 |

*pp = percentage points. The O/E framework describes within-stratum first-time PI representation in the disrupted versus non-disrupted cohort and is a descriptive complement to the adjusted Poisson regression (see Results).*

The structural divergence between Track A (null) and Track B (substantially elevated) is a substantive finding, not a measurement artifact. It indicates that the relevant exposure is the PI’s index-time activity status, that is, whether the PI was actively early-career at the calendar moment of disruption, rather than the PI’s start-time status, that is, whether the disrupted project was the PI’s debut grant. We return to this interpretation in the Discussion.

### Fiscal year 2024 common-anchor analysis

To address the asymmetry whereby disrupted projects index at their latest fiscal year in window (predominantly 2025) while non-disrupted projects index at their first fiscal year (predominantly 2024), we restricted both groups to fiscal year 2024 index records (n=63,011, of which 1,416 were disrupted) and re-estimated the primary Poisson model. Under this common-anchor identification, the Track B absolute first-time effect attenuated to RR 1.22 (0.95 to 1.58, p=0.12), while the mechanism-first effect persisted at RR 1.48 (1.07 to 2.06, p=0.019). These results indicate that part of the elevated risk in the primary analysis reflects calendar-time confounding, but that the mechanism-first signal remains statistically significant under the most conservative identification strategy.

### Sensitivity analyses

Across the nine pre-specified sensitivity analyses for the Track B absolute first-time exposure, point estimates ranged from RR 1.47 to 2.05, with all confidence intervals excluding the null except S2 (R01 only, n=31,864, RR 1.47, 0.71 to 3.06), which was non-significant, likely due to reduced sample size. The strongest effect was observed in S4a (excluding reinstated grants, RR 2.05, 1.51 to 2.79), suggesting that the inclusion of subsequently reinstated grants attenuates the primary effect. The strict and lenient PI linkage definitions yielded identical estimates (S5: RR 1.77 in both), confirming that the result is robust to the PI identifier construction. Figure 4 and Table 3 display the full sensitivity profile.

**Figure 4.**
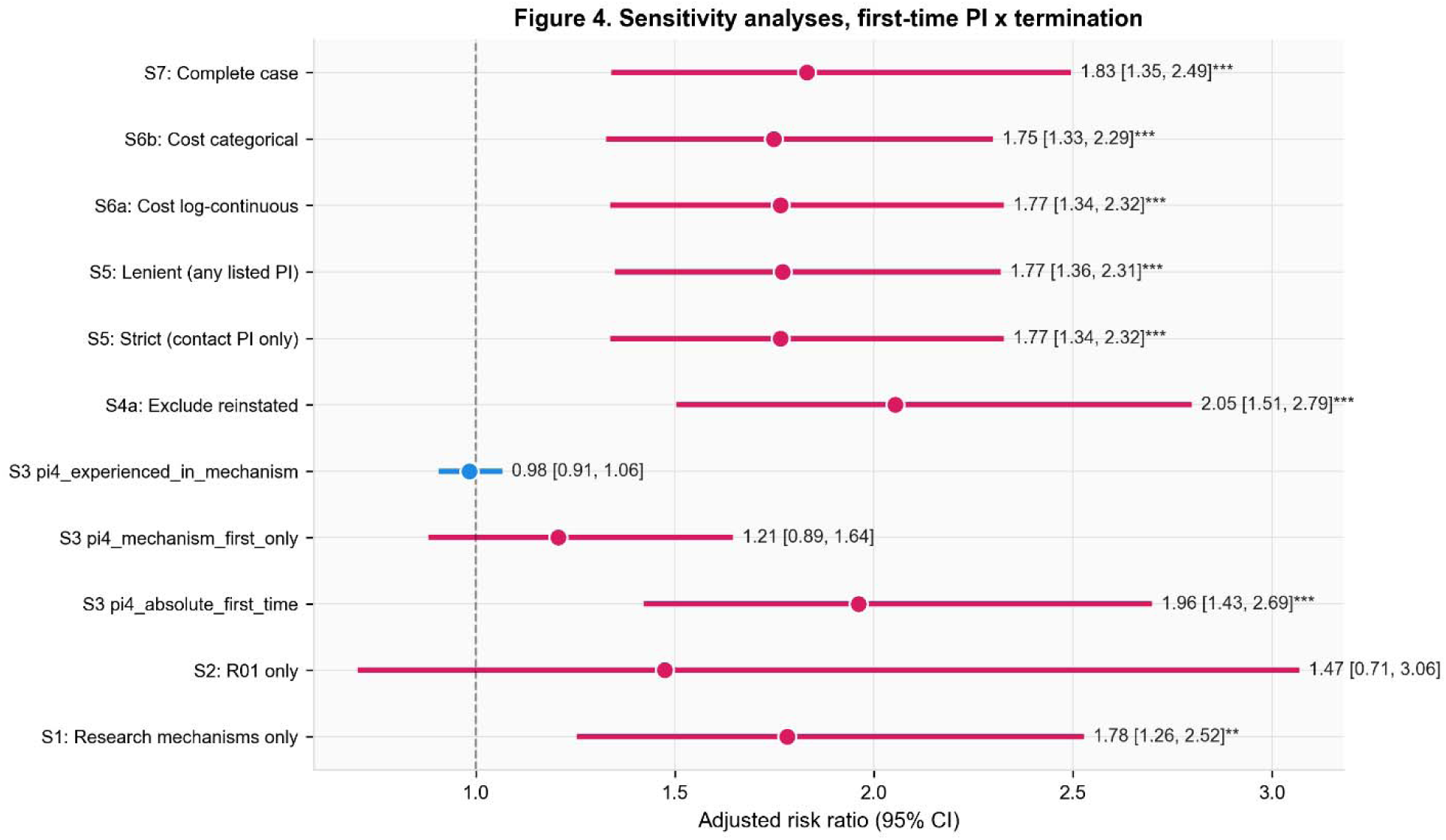
Sensitivity forest plot for Track B absolute first-time PI. Adjusted relative risks (95% CIs) for Track B absolute first-time PI across the primary analysis and nine pre-specified sensitivity analyses. The primary estimate is RR 1.77 (1.34 to 2.32). Most analyses yield a statistically significant elevated effect.

**Table 3.** Sensitivity analysis summary (Track B absolute first-time exposure) Adjusted relative risks from the modified Poisson regression for Track B absolute first-time PI across the primary analysis and nine pre-specified sensitivity analyses.

| Analysis | n | RR [95% CI] | p-value |
| --- | --- | --- | --- |
| Primary (full-cohort Poisson) | 80,976 | 1.77 [1.34, 2.32] | 4.5×10<sup>-</sup>□ |
| S1 Research mechanisms only | 60,328 | 1.78 [1.26, 2.52] | 0.001 |
| S2 R01 only | 31,864 | 1.47 [0.71, 3.06] | 0.30 |
| S3 4-level (vs experienced_all) | 80,976 | 1.96 [1.43, 2.69] | 3.1×10<sup>-</sup>□ |
| S4a Exclude reinstated | 79,003 | 2.05 [1.51, 2.79] | 4.4×10<sup>-</sup>□ |
| S5 Strict PI linkage | 80,976 | 1.77 [1.34, 2.32] | 4.5×10<sup>-</sup>□ |
| S5 Lenient PI linkage | 80,976 | 1.77 [1.36, 2.31] | 2.7×10<sup>-</sup>□ |
| S6a Cost log-continuous | 80,976 | 1.77 [1.34, 2.32] | 4.5×10<sup>-</sup>□ |
| S6b Cost categorical | 80,976 | 1.75 [1.33, 2.29] | 5.4×10<sup>-</sup>□ |
| S7 Complete case | 58,255 | 1.83 [1.35, 2.49] | 1.1×10<sup>-</sup>□ |
| FY2024 common-anchor (absolute) | 63,011 | 1.22 [0.95, 1.58] | 0.12 |
| FY2024 common-anchor (mechanism) | 63,011 | 1.48 [1.07, 2.06] | 0.019 |

*RR = relative risk. FY2024 = fiscal year 2024.*

### Subgroup interactions

By mechanism category (Figure 5, bottom), the signal was concentrated in research-mechanism grants (R, U, P, D, S codes: RR 1.78, 1.26 to 2.52, n=60,328). Training and fellowship mechanisms (F, T) yielded a directionally protective but non-significant estimate (RR 0.69, 0.45 to 1.07, p=0.10), and career development (K codes) was null (RR 0.98, 0.52 to 1.82, p=0.94).

**Figure 5.**
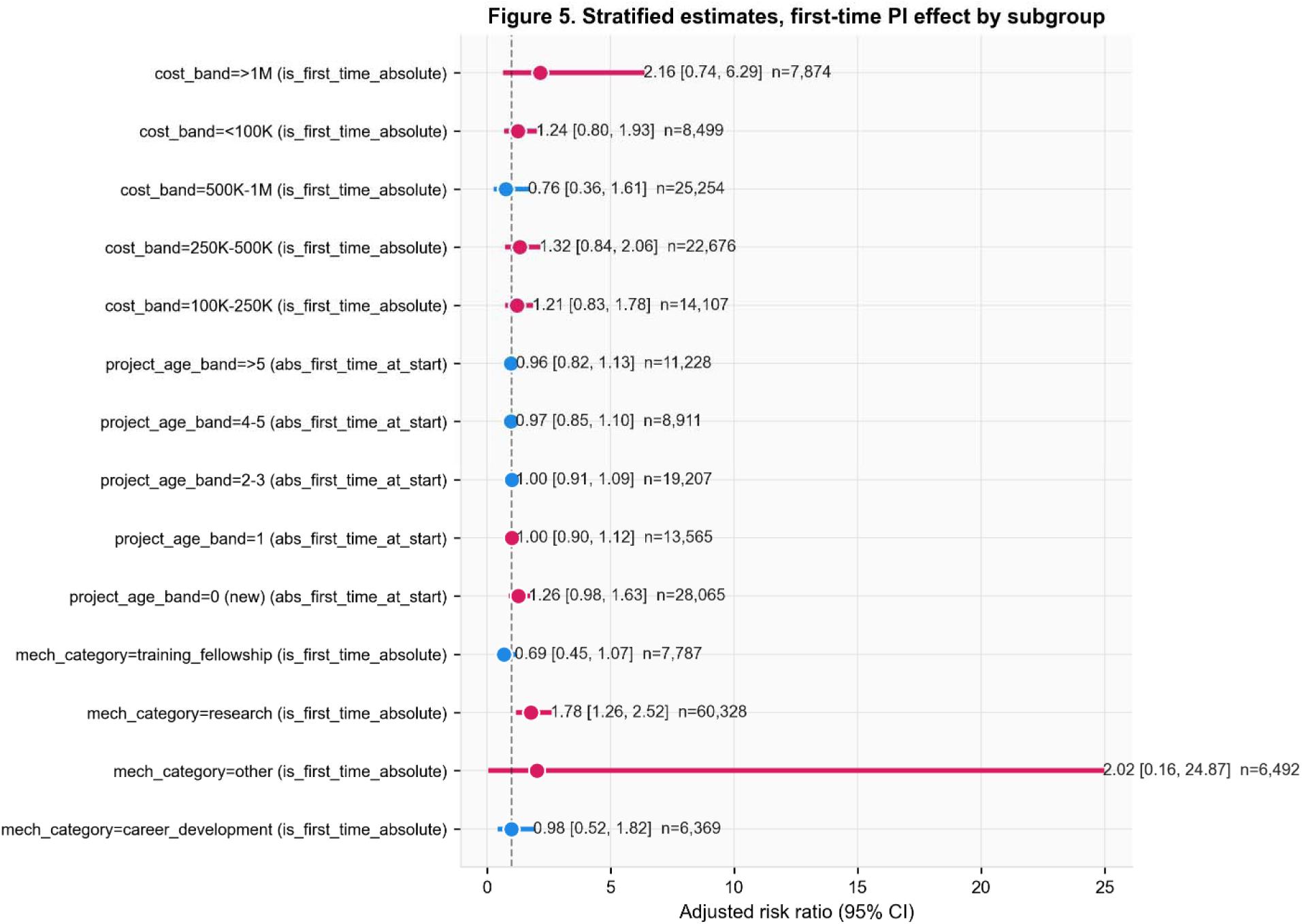
Subgroup interaction forest plots. Forest plot showing pre-specified subgroup interactions. Bottom: by mechanism category (Track B exposure); the signal is concentrated in research-mechanism grants (R/U/P/D/S codes). Middle: by project age band (Track A exposure); the Track A effect is concentrated entirely in newly started (age 0) projects, with all older age bands at or below unity. Top: by cost band (Track B exposure); all point estimates are directionally consistent with the headline, but each subgroup is underpowered for individual significance. Subgroups with fewer than 50 disrupted grants are excluded from the visualization.

By project age band at index (Figure 5, middle), the Track A start-time effect was concentrated entirely in newly started projects (age 0, RR 1.26, 0.98 to 1.63, p=0.075). For all older age bands, the Track A relative risk was at or below unity (RR 1.00, 1.00, 0.97, and 0.96 for ages 1, 2 to 3, 4 to 5, and greater than 5, respectively). This pattern is the structural empirical evidence underlying the divergence between Track A and Track B: the apparent Track A signal is concentrated in newly started projects, where the two tracks coincide, and disappears as soon as project age advances.

By cost band (Figure 5, top), all point estimates were directionally consistent with the headline finding, but each subgroup was underpowered for individual statistical significance.

### Reinstatement asymmetry

Among the 4,961 disrupted grants, those eventually flagged as reinstated had a lower prevalence of first-time PIs than those that remained un-reinstated. Reinstated grants comprised 3.85% absolute first-time PIs (76 of 1,973), compared with 5.39% in not-reinstated grants (161 of 2,988). The corresponding proportions for mechanism-first PIs were 6.74% versus 8.80%. The absolute differences are modest, but the direction is consistent with the primary finding: first-time PIs were both more likely to be disrupted and slightly less likely to be subsequently restored.

### Burden

The gross burden of the disruption wave on Track B absolute first-time PIs was 237 grants with a total committed award value of approximately $11.6 million, and 396 mechanism-first grants with a corresponding cost of approximately $59.3 million. Under the Track A start-time framing, 1,495 absolute first-time PIs and 3,330 mechanism-first PIs were affected. After adjustment, applying the attributable-fraction formula to the primary RR of 1.77 yields approximately 103 of the 237 Track B absolute first-time PI disruptions (43.5%) attributable to first-time status, corresponding to approximately $5.0 million in adjusted-attributable disruption among the most vulnerable career-stage group.

## Discussion

### Principal findings

In a retrospective cohort study of 80,976 active NIH research grants in fiscal years 2024 to 2025, first-time and mechanism-first principal investigators faced disproportionately elevated risk of administrative disruption during the wave that peaked in May 2025. After adjustment for project mechanism, NIH institute, cost, project age, support year, application type, institution type, and US region, absolute first-time PIs faced a 77% elevated relative risk of disruption (adjusted RR 1.77, 1.34 to 2.32). Mechanism-first PIs faced a 57% elevated risk (RR 1.57, 1.16 to 2.11). The effect was concentrated in research-mechanism grants (RR 1.78), was robust across eight of nine pre-specified sensitivity analyses, and was confirmed under a more conservative fiscal year 2024 common-anchor analysis for mechanism-first PIs (RR 1.48, 1.07 to 2.06).

The most informative secondary finding is the structural divergence between two operationally distinct first-time PI constructs. The start-time (Track A) framing, which asks whether the disrupted project was the PI’s debut grant, yielded null adjusted estimates (RR 0.98 for absolute first-time). The index-time (Track B) framing, which asks whether the PI had any prior NIH grant at the calendar moment of disruption, yielded the substantial elevated effect. When stratified by project age band, the Track A effect was concentrated entirely in newly started projects and vanished for projects in their second year of support or later. This pattern indicates that the relevant exposure is not whether a PI was new at project initiation, but whether they were currently early-career at the moment administrative action occurred.

### Strengths and limitations

The principal strength of this study is the comprehensive cohort framing. The full population of NIH active research grants in the analytic window served as the denominator, and the disrupted cohort was defined inclusively across all five disruption status categories present in the source registry, avoiding selection on the narrowest “terminated” definition alone. PI history covered all fiscal years present in the NIH RePORTER data, allowing precise calendar-time anchoring of first-time PI status. The primary estimator was a modified Poisson regression with institution-clustered standard errors, the established choice for binary outcomes with substantial adjustment [14,15]. The nine pre-specified sensitivity analyses and three subgroup interactions provide an extensive characterization of the effect’s robustness and heterogeneity.

The principal limitation is the structural asymmetry between the index fiscal years of the two groups. Disrupted projects were indexed at the calendar moment of disruption (predominantly 2025), and non-disrupted projects were indexed at the start of their active period (predominantly 2024). This is the conventional cohort design for an event-time exposure of this kind, but it introduces a calendar-time channel through which the relative risk can be partly inflated by the disproportionate concentration of disruptions in late fiscal year 2025. We addressed this by conducting a pre-specified fiscal year 2024 common-anchor analysis, restricting both groups to 2024 index records. Under this more conservative identification, the mechanism-first effect persists (RR 1.48, 1.07 to 2.06) while the absolute first-time effect attenuates to a non-significant RR 1.22 (0.95 to 1.58). The true causal magnitude of the absolute first-time effect likely lies between the primary estimate and the common-anchor estimate; the mechanism-first effect appears more robust to this concern.

Second, the analyses are observational and use administrative data. Unmeasured confounders, particularly intra-institutional grant-management practices, pre-disruption performance metrics, institutional willingness to pursue legal means to reinstate disrupted grants, and PI demographic characteristics not captured in NIH RePORTER, remain unadjusted. The findings establish an association after extensive covariate adjustment, but cannot establish a causal effect of first-time PI status in the counterfactual sense.

Third, the source registry of disrupted grants depends on academic monitoring efforts and public disclosure of administrative actions; some disruptions may not yet be captured, and reinstatement statuses are evolving in real time. Our coalesced disruption date covers 99.9% of disrupted fiscal year rows in the analytic cohort, but the cohort definition itself depends on the completeness of the source registry.

### Comparison with previous work

Oliveira and colleagues provided the only prior demographic-group analysis of the 2025 wave, finding that early-career investigators, assistant professors, postdoctoral scholars, trainees, and graduate students, as well as women, were descriptively over-represented among those whose grants were cancelled [4]. Their analysis covered 2,291 grants from an earlier snapshot, did not produce adjusted relative-risk estimates against the full NIH active-grant denominator, and did not address the structural confounding between PI experience and project lifecycle stage. Our analysis extends and complements theirs in three ways: a larger and more recent cohort, an adjusted modified-Poisson framework with institution-clustered inference, and an explicit Track A versus Track B exposure framing that disentangles start-time from index-time first-time PI status. Descriptive characterisations of the wave have noted geographic concentration in the Northeast, the over-representation of certain research areas, and the heavy involvement of large research universities [1–5], but no prior peer-reviewed analysis has provided adjusted relative-risk estimates of the career-stage dimension against the full NIH active-grant denominator.

Several peer-reviewed analyses have provided complementary characterisations of the same wave from different vantage points. Liu and colleagues delivered a descriptive characterization of the terminated grants [20]. Patel and colleagues estimated that 3.5% of NIH-funded interventional trials lost grant support during the first six months of the wave, affecting more than 74,000 enrolled participants and raising direct concerns about scientific waste, data integrity, and participant safety [3]. Miller and colleagues found that 41 of 64 gender-affirming care grants were terminated within a single three-week window, with nearly $22 million of committed funding left unspent [2]. Chan and colleagues described institute-level concentration within the National Institute of Allergy and Infectious Diseases, with implications for pandemic preparedness and HIV prevention infrastructure [5]. Jaramillo and Harkness mapped 405 F31 predoctoral fellowship terminations and identified disproportionate impact on F31-Diversity awards [21,22]. Independent reporting in Nature [23] and Science [24] documented the broader trajectory of the wave and projected billions of dollars in cumulative scientific waste. Our analysis complements these characterisations by adding the first adjusted relative-risk estimate of differential disruption by PI career stage.

A separate literature on NIH workforce dynamics has documented persistent challenges for early-career investigators in the broader funding environment, including longer time-to-first-R01, age-related shifts in award distributions, and disparities by race, ethnicity, and gender [7,25–27]. Our findings extend this literature by demonstrating that an acute administrative shock can also fall disproportionately on early-career PIs, and by introducing an explicit framework for distinguishing index-time from start-time first-time PI exposures.

The methodological contribution of this paper is the explicit construction and comparison of these two distinct first-time PI constructs. Our finding that these constructs yield substantively different effect estimates in the presence of lifecycle confounding suggests that future workforce analyses should be explicit about which construct they are operationalizing and should report sensitivity to the choice.

### Implications

The elevated risk of disruption for first-time and mechanism-first PIs is concentrated in research-mechanism grants, the awards that are most directly tied to a PI’s ability to sustain a laboratory and progress along the NIH funding ladder. Training and career development grants (F, T, K codes) did not show the same effect, consistent with the institutional protections these mechanisms ordinarily carry. The substantive implication is that the wave fell disproportionately on the most precarious population within the biomedical research workforce: investigators who have transitioned from training to independent research, who depend on a single grant to firmly establish and sustain a program, and who lack the institutional buffers of established laboratories [6–8,12,28].

These findings should be situated within a broader documented pattern of disruption to the US biomedical research enterprise. Commentaries have framed the 2025 wave as a stress test of the postwar US research compact, with downstream consequences including disrupted research participants, layoffs and self-censorship among scientists, scarring of early-career trajectories, and institutional capacity loss [9,29,30]. Survey-based evidence from international radiology researchers documents perceived negative effects on US-based funding, collaboration, and motivation, with a near-majority anticipating slower research progress [31]. The disproportionate burden on first-time and mechanism-first PIs that we describe here is one specific facet of this broader systemic disruption.

The reinstatement asymmetry, where first-time PIs were both more likely to be disrupted and slightly less likely to be subsequently restored, compounds this equity concern. Although absolute differences in reinstatement rates are modest, the direction is consistent with the primary finding, and the cumulative effect on the next generation of biomedical researchers could be substantial.

For policy, the findings suggest that future administrative actions of comparable scale should be evaluated against their differential career-stage burden. Existing NIH protections for Early Stage Investigators are framed for peer review of new applications [13] and do not extend to the administrative protection of already-awarded grants. The 2025 wave demonstrates that this protective infrastructure is incomplete and that an acute administrative shock can re-concentrate workforce attrition on the most vulnerable career stage in a way that pre-award protections do not anticipate. Sustaining the long-arc productivity of US biomedical research depends on the regular formation and retention of new independent investigators [32], and any policy response to comparable future episodes should explicitly include mid-stream protections for newly funded PIs within their first 24 months of independent support.

## Conclusion

The 2025 NIH grant disruption wave fell disproportionately on first-time and mechanism-first principal investigators, with adjusted relative risks of 1.77 and 1.57, respectively, in the primary analysis and confirmed 1.48 for mechanism-first PIs under a year-confounding-free identification strategy. The effect was concentrated in research-mechanism grants, was robust across 8 of 9 prespecified sensitivity analyses, and reflects a structural divergence between index-time and start-time exposure constructs that future workforce analyses should explicitly address.

## Contributors

FA conceived the study, designed the analyses, conducted the data linkage and statistical analyses, drafted the manuscript, and is the guarantor. BM contributed to the interpretation of results and the critical revision of the manuscript. Both authors approved the final version.

## Patient and public involvement

No patients or members of the public were involved in the design, conduct, or reporting of this study, which used publicly available administrative grant data only.

## Ethical approval

This study used publicly available administrative data and a publicly curated registry; no individually identifiable personal data were used. Institutional review board approval was not required.

## Data sharing

All data used in this study are publicly available.

## Transparency

The lead author (the manuscript’s guarantor) affirms that this manuscript is an honest, accurate, and transparent account of the study being reported; that no important aspects of the study have been omitted; and that any discrepancies from the study as planned have been explained.

## Data Availability

All data used in this study are publicly available.

